# Dolichocolon as a Potential Modifier of Pediatric Ulcerative Colitis Phenotype

**DOI:** 10.1101/2025.08.20.25333704

**Authors:** Richard Kellermayer, Donovan P. Berens, Réka G. Szigeti

**Author notes:** Address correspondence to: Richard Kellermayer; Section of Pediatric Gastroenterology, Hepatology & Nutrition, Baylor College of Medicine; 6621 Fannin St., CC1010.00; Houston, TX 77030-2399; Voice.

## Abstract

**Background:** Dolichocolon (DC) is an underrecognized anatomic variant associated with constipation; its association with pediatric ulcerative colitis (UC) is unknown.

**Methods:** We retrospectively reviewed abdominal MRI and CT scans in pediatric UC, Crohn’s disease (CD), and non-IBD controls, classifying DC subtypes in 111 cases.

**Results:** DC prevalence was higher in UC than CD or controls. Type 1 DC predominated in proctitis/left-sided UC (E1/E2), while Type 2 DC was enriched in extensive/pancolitis (E3/E4). Complex DC (Types 1+2) was observed only in UC.

**Discussion:** DC may modify UC distribution independent of constipation, representing a potential developmental modifier of pediatric UC phenotype and a nidus for prevention.

**Graphic abstract:** Created with BioRender by D. Berens

## Introduction

Dolichocolon (DC), an underrecognized anatomical variant characterized by redundancy of the large intestine, is strongly associated with functional constipation in both pediatric [1,2] and adult [3] populations, but emerging observations [4] link it with other common gastrointestinal disorders as well. In adults, DC has been subclassified into three types: cranial displacement of the sigmoid loop relative to the iliac crests (Type 1), caudal displacement of the transverse colon below the iliac crests (Type 2), and redundant loops at the hepatic or splenic flexures (Type 3) [3]. Due to ambiguity in defining Type 3 DC, we have historically limited diagnostic consideration in pediatric cases to Types 1 and 2. Our recent findings [1] support previous observations [5] suggesting DC is typically congenital in children, whereas it may also develop secondarily in older adults with increasing colonic stiffness and longstanding constipation.

A recent study [6] found that pediatric patients with ulcerative colitis (UC), particularly those with proctitis (E1) or left-sided colitis (E2), had a higher prevalence of constipation compared to those with extensive (E3) or pancolitis (E4) (Fisher’s exact test [FE] p = 0.0116), or Crohn’s disease (CD) (FE p = 0.0272), aligning with our own clinical observations. Given the association between DC and pediatric constipation, we examined whether DC may contribute to the pathogenesis or localization of UC in children.

## Methods

### IRB/IACUC Approval

The study was performed under institutional IRB-approved protocols (H-43969, H-50062) of Baylor College of Medicine.

### Description of Participants

We identified 39 pediatric patients with E1/E2 UC (Paris classification [7]) who had undergone diagnostic magnetic resonance enterography (MRE) and exhibited no disease progression towards extensive UC for at least six months. Of these, 22 (56 %) had a documented history of constipation, compared to 7 of 27 (26 %) in the E3/E4 UC group (p = 0.0226). To further assess the potential association of DC with UC, we compared DC prevalence (including subtype distribution) across non-constipated patients with E1/E2 UC, E3/E4 UC, L3 Crohn’s disease, and non-IBD (without inflammatory bowel disease [IBD] or other documented chronic disorder) trauma controls with normal intestinal anatomy on CT by report (Table 1). DC prevalence was analyzed as a dichotomous variable using Fisher’s exact (FE) test (GraphPad Prism, Dotmatics, Boston, MA, USA). Group sizes were determined arbitrarily given the exploratory nature of this discovery study. Statistical significance was defined as p < 0.05.

**Table 1.** Dolichocolon (DC) prevalence with subtype distribution between the patient groups examined. CD: Crohn’s disease, Ctrl: otherwise healthy, blunt force abdominal trauma patients (these patients had CT with contrast performed as trans-sectional abdominal imaging as opposed to the other groups who had MRI enterography performed as standard of care), N: number of patients, NC: no history of constipation, stdev: standard deviation. Age is in years of age when the trans-sectional imaging was performed. Italic and bold: p ≤ 0.05; italic p < 0.1.

| Patient cohorts | E1/E2 UC (NC)<sup>A</sup> | E3/E4 UC (NC)<sup>B</sup> | L3 CD (NC)<sup>C</sup> | Ctrl (NC)<sup>D</sup> |
| --- | --- | --- | --- | --- |
| N | 17 | 20 | 20 | 25 |
| Age (mean); +/-stdev | 14.6; +/- 2.2 | 14.2; +/- 2.3 | 12.7; +/- 2.2 | 12.6; +/- 3 |
| DC* (% of N) | **11 (65)<sup>C,D</sup>** | **10 (50)<sup>C,D</sup>** | 3 (15) | 4 (16) |
| Type 1 only | **7 (41)<sup>B,C,#</sup>** | 2 (10) | 2 (10) | 3 (12) |
| Type 2 only | 3 (18) | **5 (25)<sup>\*</sup>** | 1 (5) | 1 (4) |
| Type 1 and 2 | 1 (6) | 3 (15) | 0 | 0 |
| Type 1 alone and with Type 2 | **8 (47)<sup>C,D</sup>** | 5 (25) | 2 (10) | 3 (12) |
| Type 2 alone and with Type 1 | 4 (24) | **8 (40)<sup>C,D</sup>** | 1 (5) | 1 (4) |
| Type 3 only (% of N) | 3 (18) | 5 (25) | 6 (30) | 5 (20) |

Letters in superscript designate p ≤ 0.05 between the column’s group and the letter specified group. #: p < 0.1 when group A was compared to group D. *: p < 0.1 when group B was compared to group D. P values were calculated in GraphPad Prism (Dotmatics, 225 Franklin Street, Floor 26 Boston, MA 02110, USA) by Fischer’s exact testing.

### Results

The prevalence of DC on MRE did not differ significantly between E1/E2 (69 %) and E3/E4 (52 %) groups (Figure 1A; FE p = 0.1993), nor between constipated and non-constipated E1/E2 patients (73 % vs. 65 %, p = 0.7302).

**Figure 1.**
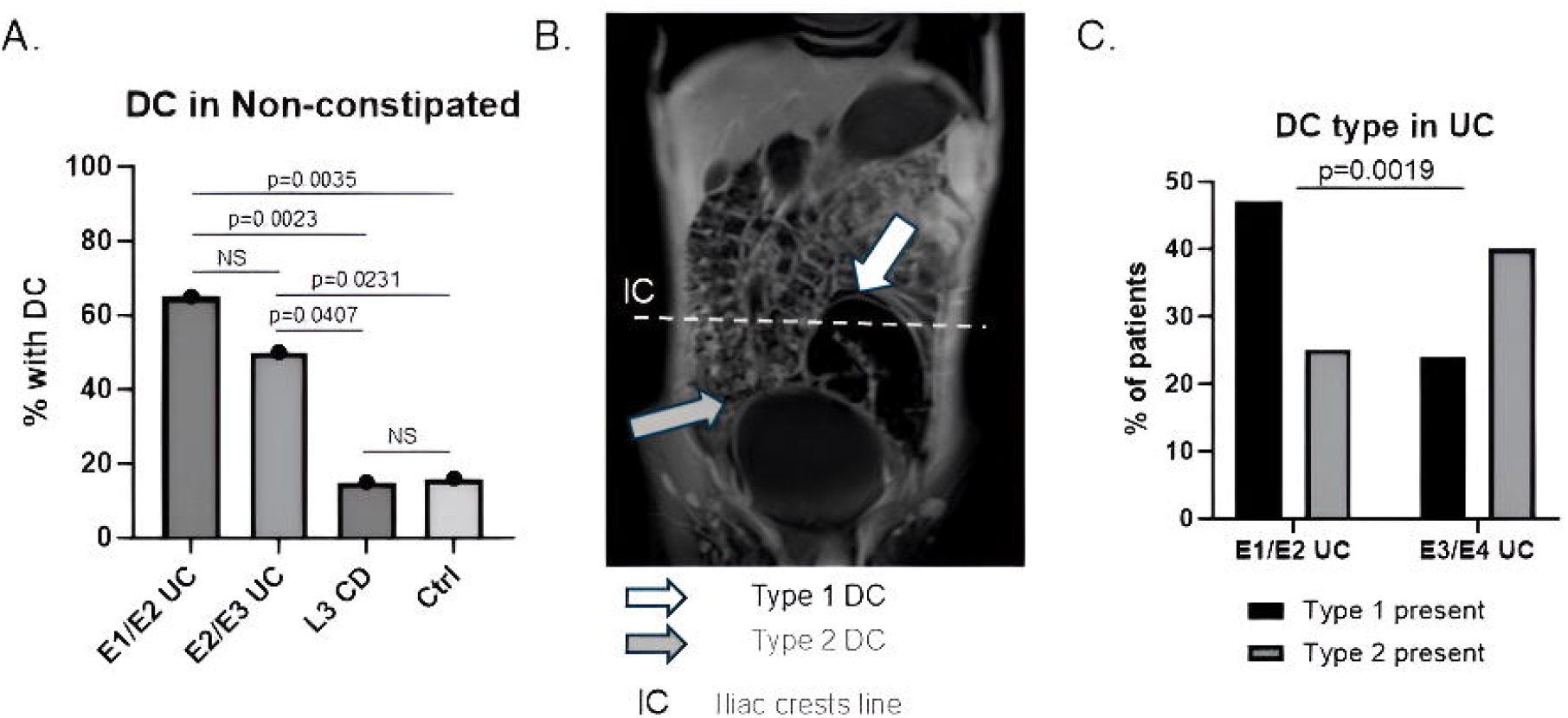
Dolichocolon (DC) may modify pediatric ulcerative colitis phenotype. **A**. DC was significantly more commonly present in E1/2 UC (n=17), and E3/4 UC (n=20), than in patients with ileocolonic (L3, n=20) Crohn’s disease (CD), or otherwise healthy patients with blunt force abdominal trauma (control: Ctrl; n=25). **B**. Complex DC (Type 1 and Type 2 simultaneously, as demonstrated in a case on the MRI enterography image) was uniquely present only in patients with UC in this cohort. **C**. The prevalence of Type 1 vs. Type 2 DC (alone or in combination) significantly separated E1/2 UC from E3/4 UC, respectively. P values demonstrated on the figures were calculated in GraphPad Prism (Dotmatics, 225 Franklin Street, Floor 26 Boston, MA 02110, USA) by Fischer’s exact testing.

DC, however, was significantly more common in both E1/E2 and E3/E4 UC groups compared to L3 CD and non-IBD controls (Figure 1A). Notably, combinations of Type 1 and Type 2 DC (Figure 1B) were observed exclusively in UC patients, with none identified in CD or healthy control cohorts. The increased prevalence of DC in E1/E2 UC was primarily attributable to Type 1 DC, whereas Type 2 DC was more prominent in the E3/E4 UC group (Figure 1C). The frequency of isolated Type 3 DC, which we excluded from primary analysis due to definitional ambiguity, was similar across all groups (Table 1).

## Discussion

Our findings suggest that DC, even in the absence of clinically significant constipation, may influence the anatomic distribution of UC. The results indicate that Type 1 DC may promote left-sided disease, while Type 2 DC may predispose to extensive or pancolitis, possibly by directing inflammatory processes to specific colonic regions (Figure 1C). As DC results in increased colonic length without corresponding increases in volume [3], it may expand epithelial surface area and augment host-microbiome interactions relevant for IBD pathogenesis [8], which may induce autoimmunity in susceptible segments. Segmental redundancy may lead to intermittent mechanical kinking and fluctuating epithelial oxygenation in the colon, potentially amplifying localized inflammation [9] and favoring a UC phenotype over CD in genetically and environmentally predisposed individuals. Recent organoid studies have shown elevated epithelial oxygen consumption and hypermetabolic stress in ulcerative colitis [10], consistent with our hypothesis that even subtle DC-associated vascular compromise could predispose to disease initiation and segmental expansion in susceptible individuals.

While UC and CD share many genetic and environmental risk factors [11], our observations are the first to implicate DC as a potential developmental modifier of UC phenotype in over 50 % of the cases. These findings may help explain not only the development of UC in the context of IBD susceptibility but also the persistence of segment-specific disease in a subset of patients. Thus, our discovery findings can set the nidus for potential interventions that may prevent the development of UC in children at increased risk for the disease, such as those with first-degree relatives suffering from IBD [12]. Early recognition of DC in such children may improve adherence to dietary, lifestyle [13], and bowel-management interventions, strategies that can optimize colonic function and potentially reduce UC risk.

Further investigation is warranted to elucidate the mechanistic and clinical implications of DC in IBD pathogenesis.

## Acknowledgements

R.K. was supported by the Gutsy Kids Fund, graciously maintained by Brock Wagner, the Klaasmeyers, the Frugonis, and other generous donor families.

## Author contributions

RK: conceptualization, acquisition and analysis of data, manuscript drafting and final submission; DPB: graphics, critical revision; RGS: conceptualization, critical revision

## Data availability statement

The datasets generated and/or analyzed during the current study are not publicly available due to institutional policies, but are available from the corresponding author on reasonable request.

## Declaration of generative AI and AI-assisted technologies in the writing process

ChatGPT was used to decrease word count and enhance readability; content was reviewed and edited by the author(s), who take full responsibility.

